# The impact of malaria-protective red blood cell polymorphisms on parasite biomass in children with severe *Plasmodium falciparum* malaria

**DOI:** 10.1101/2022.02.21.22271267

**Authors:** S Uyoga, JA Watson, P Wanjiku, JC Rop, J Makale, AW Macharia, SN Kariuki, GM Nyutu, M Shebe, M Mosobo, N Mturi, KA Rockett, CJ Woodrow, AM Dondorp, K Maitland, NJ White, TN Williams

**Affiliations:** KEMRI-Wellcome Trust Research Programme, Kilifi, Kenya; Mahidol-Oxford Research Unit, Faculty of Tropical Medicine, Mahidol University, Bangkok, Thailand; Centre for Tropical Medicine and Global Health, Nuffield Department of Medicine, University of Oxford, UK; Wellcome Trust Centre for Human Genetics, University of Oxford, Oxford, UK; Institute of Global Health Innovation, Department of Surgery and Cancer, Imperial College, London, UK

**Keywords:** Malaria, haemoglobinopathies, red blood cells, genetic protection, *Plasmodium falciparum* Histidine-Rich Protein-2, *Pf*HRP2, parasite biomass, sequestration

## Abstract

Severe falciparum malaria is a major cause of preventable child mortality in sub-Saharan Africa. The sequestration of parasitized erythrocytes in the microvasculature of vital organs is a central pathophysiological feature. The plasma concentration of the parasite protein *P. falciparum* Histidine-Rich Protein 2 (*Pf*HRP2) has diagnostic and prognostic value in severe malaria. In the current study we investigate the potential use of plasma *Pf*HRP2 and the sequestration index (the ratio of plasma *Pf*HRP2 to circulating parasites) as quantitative traits in the conduct of case-only genetic association studies of severe malaria. We demonstrate the utility of this approach using data from over 2,000 Kenyan children with severe malaria, genotyped for 14 major candidate genes that were found to be associated with protection against severe malaria in previous studies. We show that *Pf*HRP2 is a more informative quantitative trait than peripheral parasite density, and that polymorphisms in four major red cell genes (the β^S^ sickle mutation in *HBB*, the blood group mutation O in *ABO*, the α-thalassaemia mutation in *HBA*, and the Dantu blood group mutation in *GYP*) are associated with substantially lower concentrations of plasma *Pf*HRP2 at admission. Further, the effect sizes we observed were considerably larger than those relating to peripheral parasite density. An unexpected outlier was the rs1541255 A>G polymorphism in *ATP2B4* for which we saw higher plasma *Pf*HRP2 concentrations, lower parasite densities and a higher sequestration index. We provide testable hypotheses for how this might be explained in the context of this specific protective allele.

## Introduction

*Plasmodium falciparum* malaria has been the dominant cause of child mortality in tropical regions for much of the last 5000 years^1^. As a consequence, malaria has played a major part in shaping the human genome through the selection of mutations that confer a survival advantage^1^. An estimated one-quarter of the variability in the overall risk of malaria is heritable, a figure that rises to one-third for severe and complicated infections^2^.

Numerous candidate malaria protective polymorphisms have been proposed during the 70 years since the “malaria hypothesis” was first put forward by JBS Haldane, in which he suggested that the high prevalence of β-thalassaemia in the Mediterranean basin reflected natural selection by malaria^3^. Although many of these candidates have not held up to detailed scrutiny^4^ there is now strong evidence in support of some. The most convincing relate to red blood cells^5^ and are particularly plausible given the central role of the erythrocyte in the biology of malaria in humans^6^.

Through a recent case-control study, we investigated associations between mutations in a wide range of candidate genes and the risk of severe *P. falciparum* malaria in Kilifi, Kenya. We found significant evidence in support of polymorphisms in 15 genes: seven (*ABO, ATP2B4, INPP4B, G6PD, HBA, HBB* and *FREM3/GYP*) involved in red cell structure or function, seven (*ADGRL2, CAND1, RPS6KL1, GNAS, ARL14, CD40LG, IL10*) non-red blood cell protein coding genes, two of which related to immune pathways, and one non-coding RNA gene (*LOC727982*)^7^. Although all were associated with a lower risk of severe malaria, only three were associated with lower peripheral parasite densities amongst the cases. Densities were approximately five-fold lower in both heterozygotes and homozygotes for the rs334 A>T allele in *HBB*, the β^s^ mutation which encodes for the structural haemoglobin (Hb) variant HbS, in comparison to wild-type normal children, while they were marginally lower in both children with blood group O and in homozygotes for the derived allele at rs1541255 A>G in *ATP2B4*.

Peripheral parasite densities are a poor measure of total parasite loads in *P. falciparum* infections because of the time-varying proportion of parasites that are sequestered in the deep microvasculature. Although they are central to the pathophysiology of severe and complicated malaria, these parasites are not reflected in the peripheral parasite density estimation^8^. Total parasite biomass, which includes both the circulating and sequestered fractions, is better reflected by the plasma concentration of *P. falciparum* Histidine Rich Protein 2 (*Pf*HRP2)^9^, a 30 kD molecule that is released from the cytoplasm of falciparum-infected red blood cells, mainly during schizont rupture^10-12^. Plasma levels of *Pf*HRP2 have diagnostic utility in differentiating between “true” severe malaria and other severe illnesses in the presence of incidental parasitaemia^13^.

In the current study we investigate associations between the malaria-protective polymorphisms identified in our earlier study^7^ and total parasite biomass estimated through plasma concentrations of *Pf*HRP2, in a cohort of children admitted to hospital with severe *P. falciparum* malaria in Kenya. We demonstrate the utility of plasma *Pf*HRP2 and the sequestration index (the ratio of plasma *Pf*HRP2 to circulating parasites) as quantitative traits in the conduct of case-only genetic association studies of severe malaria.

## Methods

### Study design and participants

The Kenyan severe malaria cohort has been described in detail previously^7^. Cases consisted of children aged <14 years who were admitted to the high dependency ward of Kilifi County Hospital (KCH) between June 11^th^ 1999 and June 12^th^ 2008 with clinical features of severe falciparum malaria. Controls were infants aged 3–12 months who were born in the same study area as cases and were recruits to a cohort study investigating genetic susceptibility to a range of childhood diseases.

### Clinical and laboratory data

Data and samples were collected through a clinical surveillance system that has been described in detail previously^14^. Severe malaria was defined as a positive blood-film for *P. falciparum* parasites together with prostration (a Blantyre Coma Score of 3 or 4), cerebral malaria (a Blantyre Coma Score of <3), respiratory distress (abnormally deep or “Kaussmaul’s” breathing), severe anaemia (haemoglobin concentration of <5 g/dL) or any other feature of severe malaria as defined by the World Health Organization^15^. Admission blood films were stained and examined by standard methods^16^ and parasite densities recorded as either the ratio of parasites to white blood cells in thick films or the proportion of parasitised red blood cells in thin films for heavier infections. Parasite densities (the number of parasites/µl of whole blood) were then calculated with reference to data from full haematological assessments, if available, or on the assumption of a white blood cell count of 8×10^3^/µL or a red blood cell count of 5×10^6^ /µL if not.

### Genotyping

Severe malaria cases were genotyped for the rs334 SNP and for the common African form of α-thalassaemia (a 3.7kb deletion in *HBA1* and *HBA2*; -α^3.7^-thalassaemia^17^) by PCR at the KEMRI-Wellcome Trust Laboratories, Kenya, as described in detail previously^7,18,19^. Genotyping for a further 119 candidate SNPs was conducted at the Wellcome Centre for Human Genetics in Oxford, UK, by use of the Sequenom iPLEX MassARRAY platform, using DNA extracted from frozen samples of whole blood as previously described^7^. Plasma concentrations of *Pf*HRP2 were batch-analysed in Kenya by ELISA in admission samples stored at -80°C using previously published methods^20^. We focused our analysis on 14 of the 15 previously associated genes: we excluded *INPP4B* because it tags the same nearby causal Dantu *GYP* mutation as *FREM3*, which is in stronger linkage disequilibrium^21,22^.

### Statistical analysis

We explored the effects of polymorphisms in *ABO, ATP2B4, G6PD, HBA, HBB, FREM3/GYP, ADGRL2, CAND1, RPS6KL1, GNAS, ARL14, CD40LG, IL10* and *LOC727982* on three quantitative parasite-specific traits: (i) peripheral parasite density (the number of circulating parasites per µL); (ii) plasma *Pf*HRP2 concentration (ng/mL), a proxy measurement of the total parasite biomass; and (iii) the ratio of the plasma *Pf*HRP2 concentration to the circulating parasite density (ng/parasite), which is proportional to the previously reported sequestration index^9^. The previously reported calculation of the sequestration index requires multiplying the plasma *Pf*HRP2 by a constant term which is an estimate of the multiplication rate over the previous lifecycle^9^. The *Pf*HRP2 to parasite ratio is simpler to interpret as it does not make any assumptions about multiplication rates (which could potentially vary considerably across individuals) an issue that is particularly pertinent in the context of malaria-protective genes. For all three endpoints we used a log_10_ transformation as the parasite density and plasma *Pf*HRP2 concentration are approximately log-normally distributed. The *Pf*HRP2 to parasite ratio is thus expressed as the difference between the two log_10_ transformed variables. To aid the comparison of effect sizes across the three quantitative traits we standardized each dependent variable to obtain distributions with mean 0 and standard deviation 1. To account for parasite densities that were below the lower limit of detection by thick film microscopy (∼50 parasites/µL), we used a log_10_(x+50) transformation for the peripheral parasite density. We excluded non-measurable *Pf*HRP2 concentrations (22/1,522 samples) as these could represent *HRP2/3* parasite gene deletions or assay errors. Out of 2,220 genotyped severe malaria patients, measurable *Pf*HRP2 concentrations were available for 1,500 patients and parasite densities were available for 2,184 patients. In addition, we included patient data on platelet counts, admission haemoglobin and other key admission variables relating to patient severity and demographics. To increase power, we performed multiple imputation using random forests for the patients with missing *Pf*HRP2 concentrations and missing parasite densities, excluding patients who had missing *Pf*HRP2 concentration, parasite count and platelet count data (n=22 patients). 10 imputed datasets were generated (R package *missForest*).

Quantitative trait associations were assessed under linear models with Gaussian errors. All models were adjusted for self-reported ethnicity. We fitted a range of association models to the parasite density and plasma *Pf*HRP2-based traits including general two-parameter models, additive, heterozygous and recessive models, and determined the best fitting models from the model likelihoods. The latter three models of association were then compared against the general two-parameter model using a likelihood ratio test with 1 degree of freedom. A p-value threshold of 0.05 was used to reject an association model. All p-values are reported without correction for multiple testing. Parameter estimates for each imputed dataset were combined using Rubin’s rules (R package *mitools*). To demonstrate the phenotypic information contained in the plasma *Pf*HRP2 for the severe malaria cases, we estimated case-control odds-ratios for protection against severe malaria, using probabilistic data-tilting to down-weight patients who likely did not have severe malaria, using a previously reported model based on *Pf*HRP2 and platelet counts^13^. All analyses were done in R v4.0.2.

## Results

After multiple imputation, we had full data on a total of 2,198 patients for our final analyses. The key predictors of the plasma *Pf*HRP2 concentration were the admission platelet count (log transformed as previously described)^13^, the admission haemoglobin concentration, and parasite density. The overall distributions of both parasite densities and plasma *Pf*HRP2 concentrations are summarized in Figure 1 panels A and B while the results of linear quantitative trait association models between each of these parameters as well as the plasma *Pf*HRP2 to parasite density ratio and the 14 severe malaria associated genetic polymorphisms are shown in Figure 1C. We also performed a sensitivity analysis using the complete case data. The pattern of the missing data is shown in Supplementary Figure 1.

**Figure 1.**
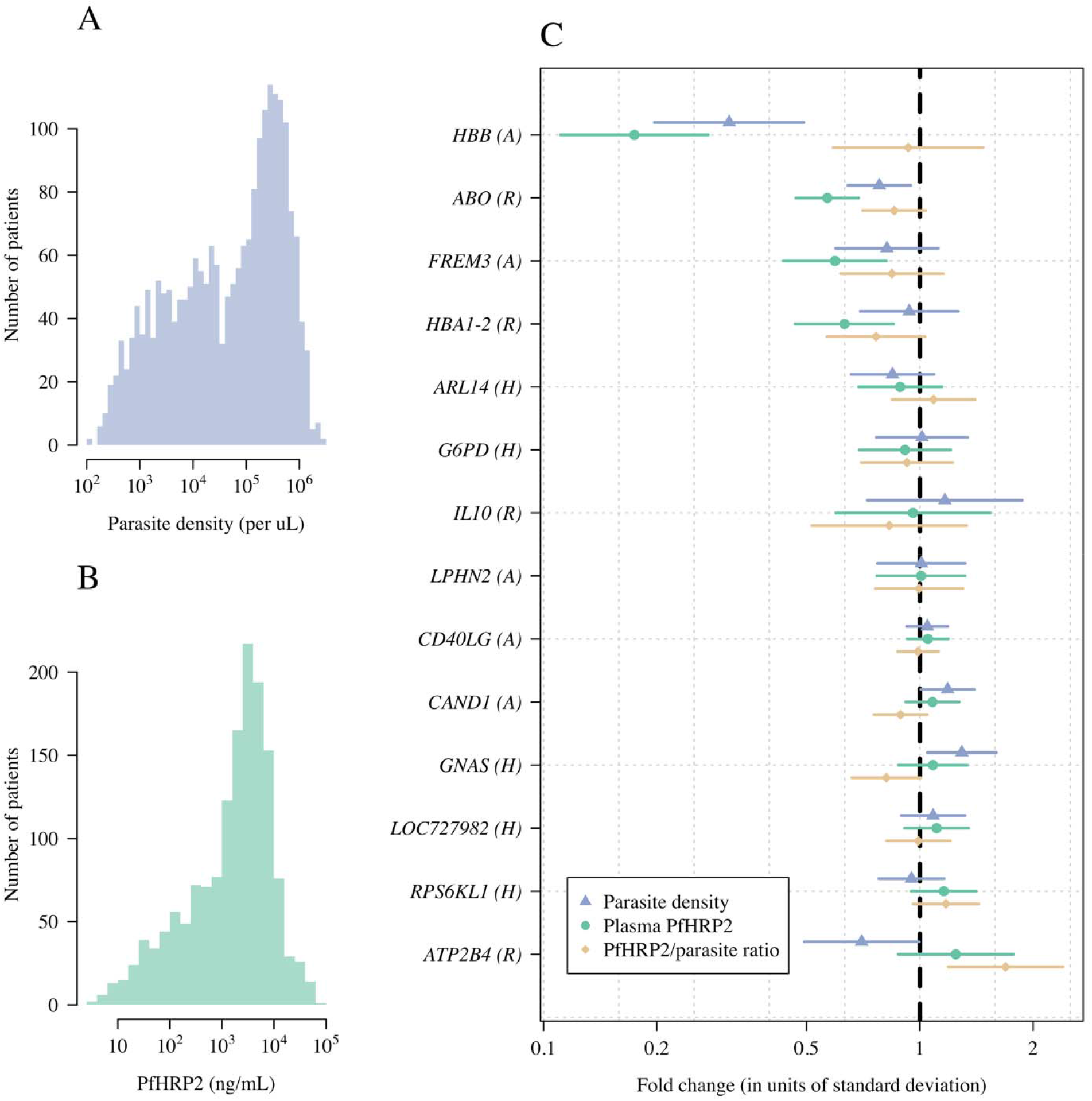
A case-only association study with 14 targeted variants using as quantitative traits the parasite density, the plasma *Pf*HRP2 concentration, and the *Pf*HRP2/parasite ratio.

As reported previously, only three polymorphisms were associated with lower parasite densities in cases (Figure 1C: *HBB*: reduction in geometric mean of 70% [95% CI 50-80]; *ATP2B4*: reduction of 30% [95% CI: 0-51]; *ABO*: reduction of 22% [95% CI: 5-36]). In comparison, four of the six major red blood cell polymorphisms (*HBB, HBA1-2, FREM3/GYP, ABO*) were associated with plasma *Pf*HRP2 concentrations, all with larger effect sizes (Figure 1C: *HBB*: reduction in geometric mean of 83% [95% CI: 73-89]; *ABO*: reduction of 43% [95% CI: 31-53]; *FREM3/GYP*: reduction of 41% [95% CI: 18-57]; *HBA*: reduction of 37% [95% CI: 15-53]). For these four polymorphisms we compared the best fitting models of association in this case-only analysis to the best fitting models of association from previously reported case-control analyses. There were notable differences with previously published case-control studies regarding the best fitting models for these major polymorphisms. For rs334 (HbS) a heterozygous model of association was reported in the largest case-control study conducted to date, which included almost 12,000 cases and more than 17,000 controls, showing no apparent protective effect of homozygous sickle cell anaemia (HbSS)^5^. However, in this case-only analysis of *Pf*HRP2 as a quantitative trait, the best fitting model of association was additive (i.e. greater protection in homozygotes than in heterozygotes), and offered strong evidence for rejecting the heterozygous model (likelihood ratio test comparing the heterozygous model with the two-parameter model: *p*=10^−5^). For *HBA1-2* (α^-3.7^-thalassaemia) the best fitting model was recessive, although the data were also consistent with an additive effect (only the heterozygous model was rejected, p=0.001). The best fitting models for the rs186873296 mutation in *FREM3* (Dantu), and the O blood group mutation in *ABO* (rs8176719) assumed additive and recessive models of association respectively, consistent with our previous study^7^. Polymorphisms in *ATP2B4, G6PD, RPS6KL1, LOC727982, ARL14, LPHN2, IL10, CAND1*, and *GNAS* were not associated with differences in baseline *Pf*HRP2 and so no best model of association could be determined.

For two key polymorphisms, the α^-3.7^ deletion in *HBA* and *FREM3* which tags the Dantu blood group, no significant associations were seen for parasite density but significant associations were seen for the plasma *Pf*HRP2 concentration (p=0.007 and p=0.002, respectively) (Figure 2). Similarly, while only a marginal association was seen with parasite density for *ABO* (p=0.01), a highly significant association was seen with plasma *Pf*HRP2 concentration (p=10^−7^, Figure 2). A notable outlier was the rs1541255 polymorphism in *ATP2B4* (Figure 3). This polymorphism was associated with the third strongest level of protection in our previous multi-country case-control analysis^23^ and was associated with marginally lower peripheral parasite densities (p=0.05). However, in the current analysis we found a trend towards higher plasma concentrations of *Pf*HRP2 (p=0.08), an effect in the opposite direction to that expected. As a result, the association with the sequestration index (the *Pf*HRP2/parasite ratio) was significant (70% increase in the geometric mean ratio [95% CI: 20-140], p=0.001). Haemoglobin concentrations at admission were no lower in homozygotes than they were in wild type or heterozygotes, as might be expected if the higher plasma PfHRP2 concentrations were a consequence of a longer duration of illness.

**Figure 2.**
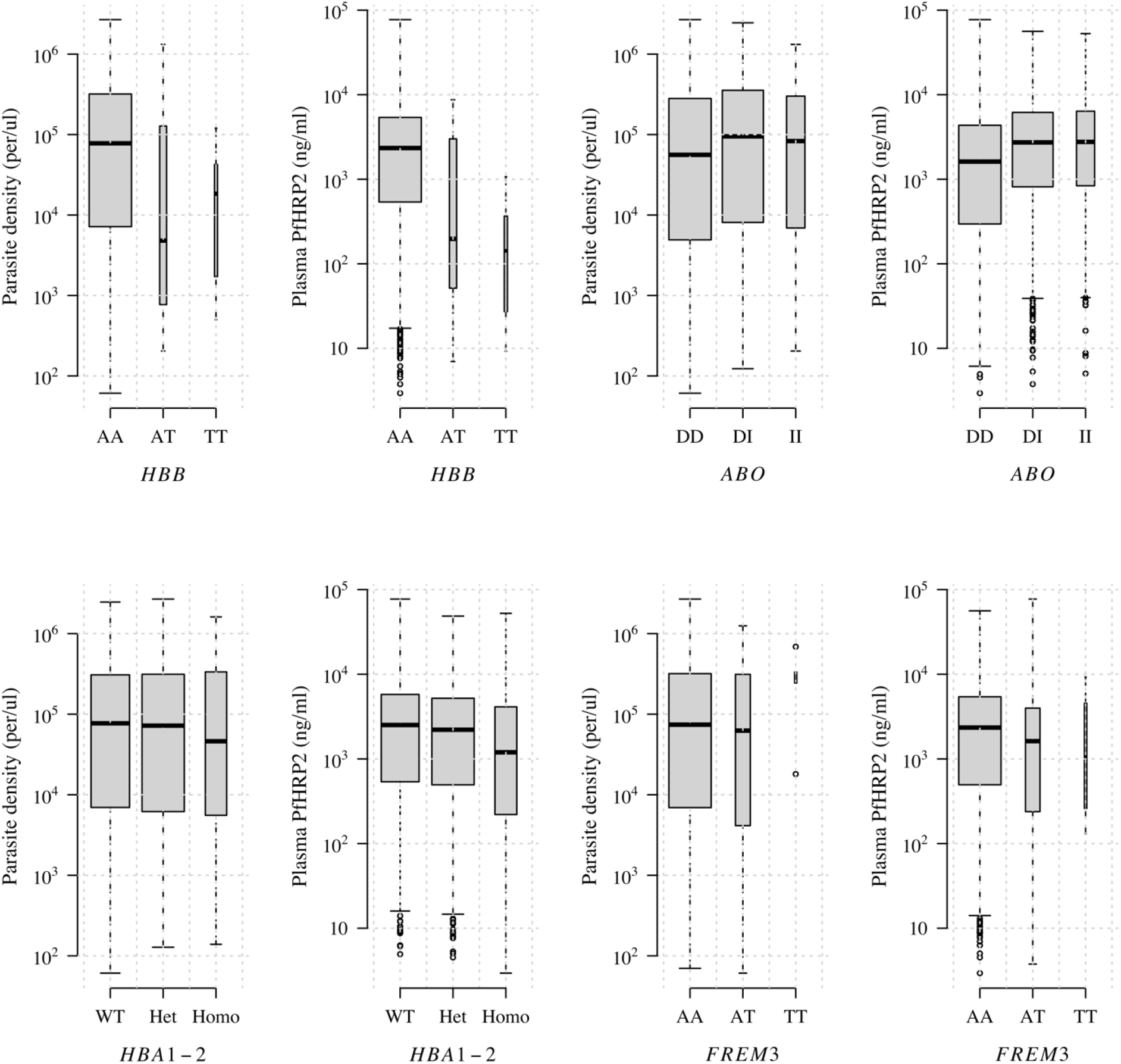
The distributions of plasma *Pf*HRP2 concentrations and parasite densities across *HBB* (rs344), *ABO* (rs8176719), *HBA1-2* (α-thalassaemia), and *FREM3/GYP* (rs186873296) genotypes. The width of each boxplot is proportional to the square root of the number of data points.

**Figure 3.**
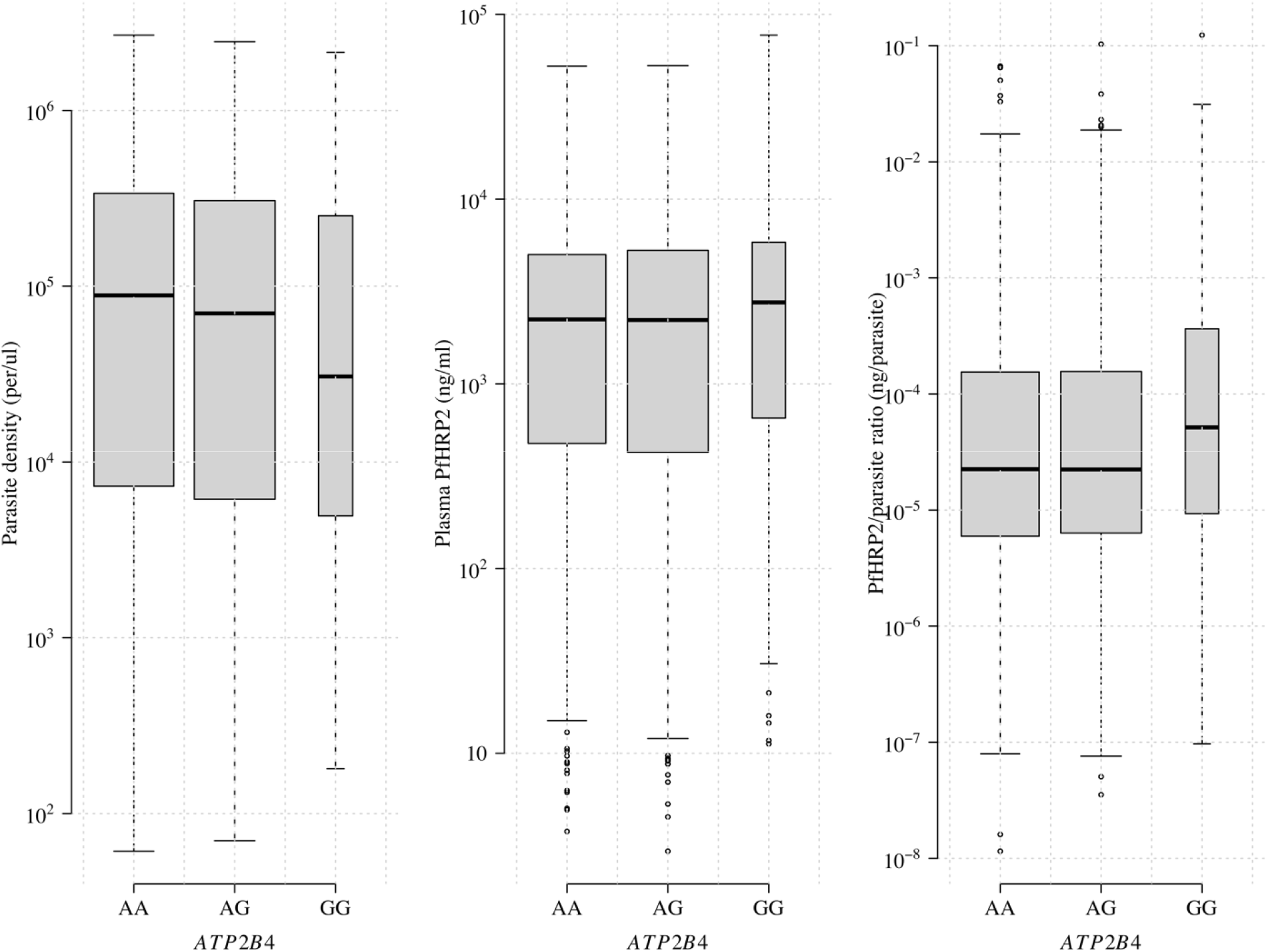
The distributions of the plasma *Pf*HRP2, the parasite density and the *Pf*HRP2/parasite ratio across the *ATP2B4* genotypes (rs1541255).

**Figure 4.**
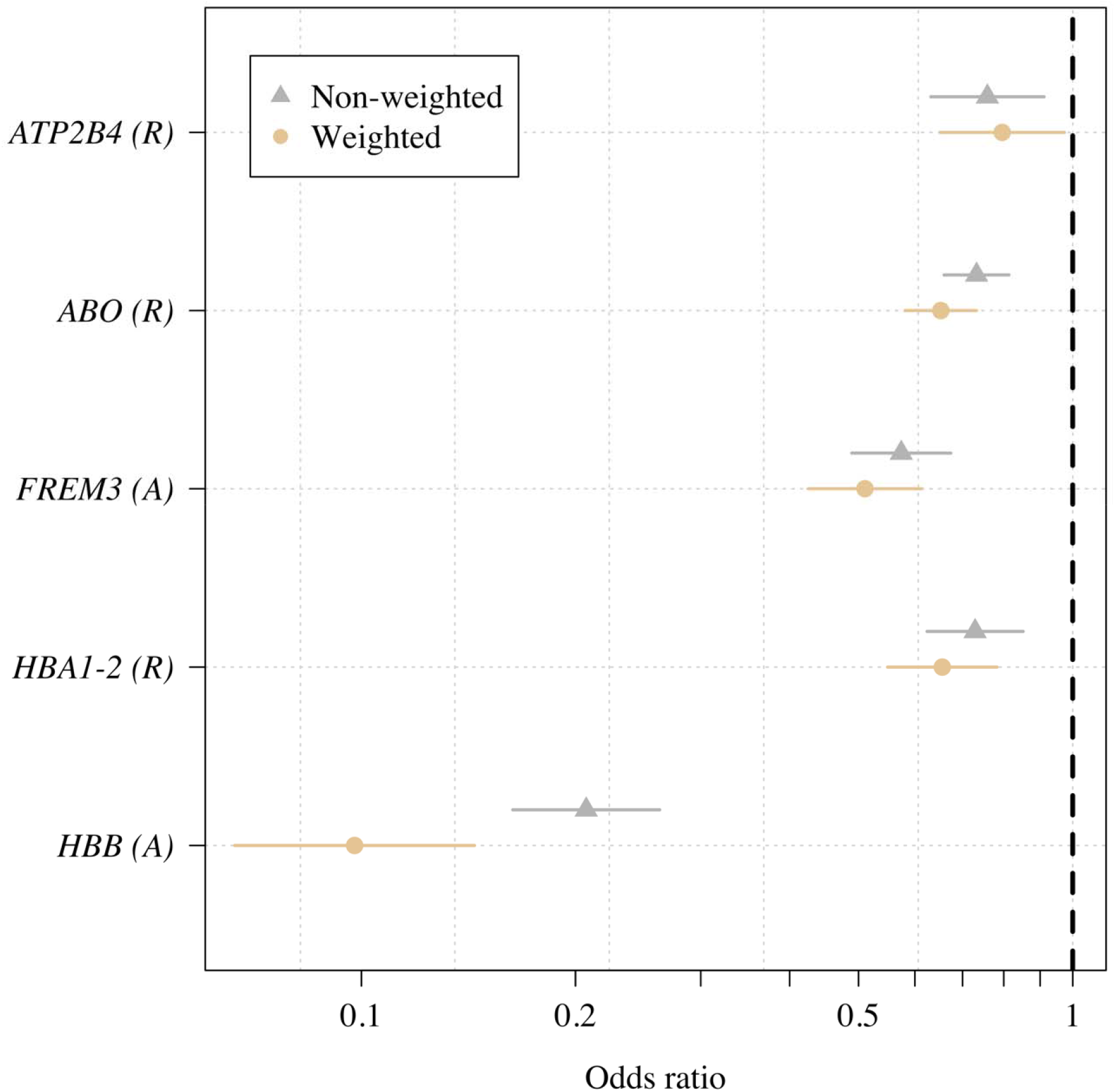
A comparison between the non-weighted (standard approach) and the weighted case-control odds-ratios for severe malaria protection for the five major red cell polymorphisms.

We re-estimated case-control odds-ratios under a data-tilting framework for our five major red blood cell genetic polymorphisms using previously published probability weights based on the plasma *Pf*HRP2 and the platelet count to probabilistically downweigh patients who were unlikely to have true severe malaria^13,24^. This increased the evidence that *HBB, ABO, HBA1-2* and *FREM3/GYP* are associated with protection against severe malaria when comparing against the non-weighted case-controls odds-ratios (for *HBA1-2*: p=10^−5^ versus p=10^−4^; *ABO*: p=10^−13^ versus p=10^−8^; *FREM3*: p=10^−12^ versus p=10^−11^). However, it decreased the evidence (p=0.02 versus p=0.003) for *ATP2B4*.

## Discussion

In this large cohort of children with severe *P. falciparum* malaria, we show that plasma *Pf*HRP2 concentration is a useful phenotypic measure that can be used to validate genetic associations discovered through case-control studies. Consistent with previous reports, plasma *Pf*HRP2 concentration is a better measure of disease severity than peripheral parasite density, and more accurately pinpoints true severe malaria. Our study shows that by providing a simple quantitative assessment of both disease severity and diagnostic accuracy, plasma *Pf*HRP2 concentration can be a useful quantitative trait for use in genetic association studies.

During the period since the “malaria hypothesis” was first put forward^3^ epidemiological support has been gathered with regard to a number candidate genes. However, in most cases the mechanisms involved remain incompletely understood^4^. For example, several different hypotheses have been put forward regarding the best-established candidate, the sickle mutation in *HBB*. The mechanisms proposed have fallen into three broad categories. First, the early *in vitro* studies suggested that HbS containing red blood cells are less supportive of *P. falciparum* parasite growth and development, particularly under conditions of low oxygen tension^25-27^. Second, there is evidence that HbS attenuates the pathophysiological consequences of malaria through mechanisms that include reduced intracellular transport and expression of surface proteins that lead to the sequestration of late-stage parasite-infected red cells on the vascular endothelium^28-30^. Finally, it has also been suggested that malaria parasite-infected HbS containing red blood cells might be removed more rapidly from circulation through immunological mechanisms, particularly involving the spleen^31-33^. Our observation that both peripheral parasite densities and plasma *Pf*HRP2 are considerably lower in children carrying the β^s^ mutation is broadly consistent with all these mechanisms. Similar mechanisms have been proposed regarding to the malaria-protective effect of α-thalassaemia yet, as in our current study, no associations have been found with peripheral parasite densities in the majority of epidemiological studies^4^. Our novel finding that plasma *Pf*HRP2 concentrations are substantially lower in children with α-thalassaemia supports the conclusion that *P. falciparum* parasites are less able to thrive in carriers of this condition than they are in normal children. Finally, in a recent mechanistic study we found that red cells expressing the rare Dantu blood group are resistant to invasion by *P. falciparum* merozoites because they are more tense than normal cells^22^. While in the current study this was not reflected by reduced parasite densities, we did see a 40% reduction in *Pf*HRP2 concentration in Dantu children overall. Furthermore, in keeping with the biological data on red cell invasion^22^, the best fitting model was additive, providing epidemiological support for the proposed biological mechanism.

One unexpected finding of our study was the relationship between the polymorphism in *ATP2B4* and the plasma *Pf*HRP2 concentration and the sequestration index. The polymorphism involved, which results in red cell dehydration and raised concentrations of intracellular calcium^34^, was the third strongest protective signal in the largest multi-country case-control study of severe malaria conducted to date^23^. Nevertheless, in the current study we found that although the protective allele was associated with slightly lower parasite densities, plasma concentrations of *Pf*HRP2 were unexpectedly higher as was the sequestration index (the ratio of *Pf*HRP2 to parasite density). If this corresponds to a true effect it could have several possible explanations. One would be that parasite multiplication rates are lower in *ATP2B4* homozygous patients, thus leading to higher plasma *Pf*HRP2 concentrations relative to circulating parasites. However, this would imply a longer illness course and thus should in theory be associated with a greater degree of anaemia, something we did not observe in the current study. Other potential explanations are that a greater amount of *Pf*HRP2 is released into the plasma from the red cell at schizogony because less *Pf*HRP2 is retained in the cytoadherent ruptured ghost erythrocyte, or that the mutation results in enhanced *Pf*HRP2 production. Both hypotheses are testable in laboratory experiments.

## Data Availability

All data produced in the present study are available upon reasonable request to the authors.

## Acknowledgements

This research was funded, in whole or in part, by The Wellcome Trust, Grant 093956/Z/10/C. A CC BY or equivalent licence is applied to author accepted manuscript arising from this submission, in accordance with the grant’s open access conditions. This work was conducted as part of SMAART (Severe Malaria Africa – A consortium for Research and Trials) funded by a Wellcome Collaborative Award in Science grant (209265/Z/17/Z) held in part by KM and AMD. Sample genotyping was conducted in part by the Malaria Genomic Epidemiology Network, supported by Wellcome (WT077383/Z/05/Z) and the Bill & Melinda Gates Foundation through the Foundations of the National Institutes of Health (566) as part of the Grand Challenges in Global Health Initiative. The Resource Centre for Genomic Epidemiology of Malaria is supported by Wellcome (090770/Z/09/Z; 204911/Z/16/Z). This research was supported by the Medical Research Council (G0600718; G0600230; MR/M006212/1). Wellcome also provides core awards to the Wellcome Centre for Human Genetics (203141/Z/16/Z) and the Wellcome Sanger Institute (206194). Sample collection and processing was further supported through a Programme Grant (092654) to the Kilifi Programme from Wellcome. TNW and NJW are senior and principal research fellows respectively funded by the Wellcome Trust (202800/Z/16/Z and 093956/Z/10/C, respectively). JAW is a Sir Henry Dale Fellow funded by the Wellcome Trust (223253/Z/21/Z). This paper is published with permission from the Director of the Kenya Medical Research Institute (KEMRI). No competing interests are declared.

---

**Supplementary Figure 1:**
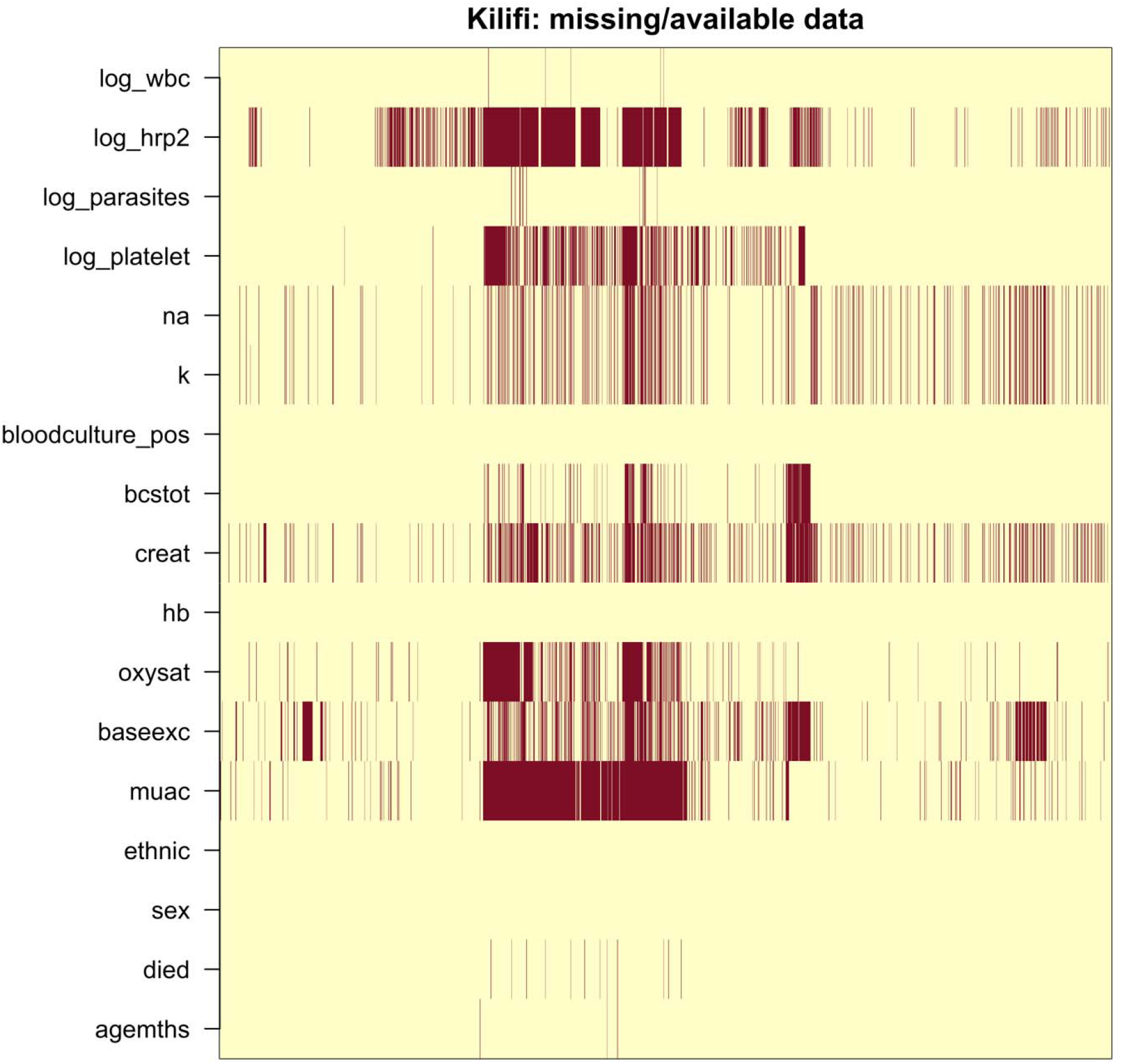
Pattern of missing data in the clinical dataset (red: missing; yellow: not missing).

